# Depression and Anxiety Symptom Networks Across the Lifespan

**DOI:** 10.1101/2025.02.17.25322400

**Authors:** Daniel Harlev, Aya Vituri, Moni Shahar, Noham Wolpe

## Abstract

**Background:** The relationship between anxiety and depressive symptoms is complex and may vary across the lifespan. Symptom network analyses offer a powerful tool to examine these interactions, but few studies have directly compared symptom networks in younger and older adults.

**Methods:** We analysed data from the Cambridge Centre for Ageing and Neuroscience (Cam-CAN) study, including 786 participants aged 18 to 88, who reported at least subclinical levels of symptoms on the Hospital Anxiety and Depression Scale (HADS). Network analysis was employed to examine symptom communities, within- and between-community connectivity, and centrality measures across age groups.

**Results:** The overall network structure, largely separating anxiety and depressive symptoms into two communities, remained stable across age groups. However, in older adults, there was a reduction in connectivity within the depression community, and a reduction in between-community connectivity for both the depression and anxiety communities. Moreover, while “panic” emerged as a consistently central symptom across both age groups, “rumination” and “restlessness” were the key bridge symptoms in young and older adults, respectively.

**Discussion:** Our findings reveal both stable and dynamic aspects of depression and anxiety symptoms across the lifespan. Reduced within-community connectivity for depressive symptoms suggests greater heterogeneity in how depression manifests in older populations. The shift in bridging symptoms, from cognitive (rumination) in young adults to somatic (restlessness) in older adults, suggests subtle yet clinically important differences in how depression and anxiety are linked across the lifespan. Our results underscore the need for tailored, age-dependent treatment in depression and anxiety.

## INTRODUCTION

Depression is a common and disabling condition, marked by the heterogeneity of its symptoms and their severity (Buch & Liston, 2021; Gutiérrez-Rojas et al., 2020). The impact of even mild “subclinical” depressive symptoms (i.e., not meeting formal diagnostic criteria) on wellbeing (Cuijpers & Smit, 2008; Noyes et al., 2022), and the substantial overlap and frequent co-occurrence of depressive and anxiety symptoms (Zbozinek et al., 2012), challenge the traditional categorical view of these conditions as distinct entities. Instead, a dimensional approach, which conceptualizes these conditions as interconnected along a spectrum of severity, may be better suited to capture their complex and intertwined nature (Vink et al., 2008).

Network analysis offers a powerful tool for examining symptom relationships within this dimensional framework (Fried et al., 2016). By visualizing and quantifying connections between symptoms, researchers and clinicians can identify central symptoms (”hubs”) that may be crucial for understanding and treating these disorders (Park & Kim, 2020). Network analysis can thus reveal critical insights into the dynamics of depression and anxiety symptoms, potentially informing more effective treatment strategies tailored to symptom heterogeneity (van Borkulo et al., 2015).

A significant contributor to symptom heterogeneity is age. The clinical presentation of symptoms of depression and anxiety differ between younger and older adults (Alexopoulos, 2019; Bergmann et al., 2023). While older adults tend to report more somatic symptoms, young adults predominantly report mood and interpersonal symptoms (Schaakxs et al., 2017). Network analyses can provide valuable insights into age-related symptom heterogeneity in depression and anxiety, by examining the relationship between symptoms and their statistical “structure”.

Previous studies have examined the symptom network of depression and anxiety in young and older adults, but separately. These studies have yielded mixed findings regarding the relationship between depression and anxiety symptoms, with inconsistencies in network structures and symptom centrality patterns within groups (Beard et al., 2016; Kaiser et al., 2021; Park & Kim, 2020; Yang et al., 2022; Zhang et al., 2023). For example, while some studies have found “loss of energy”, “loss of interest”, and “worthlessness” to be central in younger adults with major depressive disorder (Park & Kim, 2020), others have identified “felt sadness”, “uncontrollable worry” and “trouble relaxing” as more central in older adults (Yang et al., 2022; Zhang et al., 2023). To our knowledge, no study has directly compared the symptom networks of young and older adults.

This study aims to address this gap by directly comparing symptom networks in young and older adults, utilizing data from the general population across the lifespan through the Cambridge Centre for Aging and Neuroscience (Cam-CAN) study. We employed symptom network analysis to examine the structure and centrality of depressive and anxiety symptoms. By using a large, population-based sample and a standardized assessment tool, we aimed to provide a robust and comprehensive comparison of symptom networks across the lifespan. We hypothesized that while both young and older adults would show similar communities related to depression and anxiety symptoms, older adults would exhibit increased heterogeneity in symptom relationships, reflected in overall reduced centrality and connection strengths. We further hypothesized that the central symptoms would differ with age, with mood symptoms showing high centrality in young adults, while worrying symptoms would show higher centrality in older adults.

## METHODS

### Participants

We analysed data from the Cambridge Centre for Ageing and Neuroscience (Cam-CAN), which is a comprehensive research project investigating cognition across the lifespan (Shafto et al., 2014). Participants were recruited through sampling of primary care populations within specific geographic areas. In the UK, this method closely approximates a truly representative sample of the population, as registration with general practitioners is nearly universal. Data included in this study was from Stage 1 (CC3000). Ethical approval was granted by the local ethics committee, Cambridgeshire 2 (now East of England – Cambridge Central) Research Ethics Committee (reference: 10/H0308/50). All participants provided written informed consent prior to study enrolment. This study was conducted in accordance with the ethical principles outlined in the Declaration of Helsinki. We defined “young adults” as those aged 18-45 years and “older adults” as those aged 65 years and older.

### Measures and scales

In Stage 1 (CC3000), 2,681 participants completed a home interview that included demographic questionnaires (age, sex, education, medical history) and the Hospital Depression and anxiety Scale (HADS) (Bjelland et al., 2002), which is a 14-item self-report questionnaire with depression (HADS-D) and anxiety (HADS-A) subscales. We included participants who scored 4 or above on the HADS-D subscale, indicating the presence of at least subclinical depressive symptoms (Cuijpers & Smit, 2008; Szymkowicz et al., 2019). This decision was based on evidence suggesting that even mild depressive symptoms can have significant implications for individuals’ life (Meeks et al., 2011; Rodríguez et al., 2012), and is consistent with previous theoretical, data-based network structure approach (Belvederi Murri et al., 2020).

### Network computation

To compute the networks, we used similar methodology of previous network analysis studies that examined the structure of depressive and anxiety symptoms cross-sectionally (Beard et al., 2016), and other psychiatric symptoms including schizophrenia (Strauss et al., 2019; Wolpe et al., 2024). We constructed a network for all 14 HADS items, i.e., HA1-HA7 and HD1-HD7. For each item (‘node’), we computed its relationships (‘edges’) with all other 13 items. This was done separately for young and older adults. Relationship or edge strengths were calculated using the mutual information measure (MI), where the edge strength between items X and Y of the HADS was calculated by:

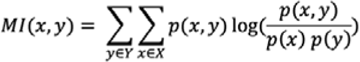

In words, MI is the sum over all possible values of X and Y of: the joint probability of X and Y, times the log of the joint probability of X and Y, divided by the product of the marginal probability of X and the marginal probability of Y.

We chose not to employ any thresholding on edge strengths to retain the full range of symptom associations, even those that might be weak or subtle, as even small connections can be clinically meaningful in understanding the complex interplay of symptoms (Borsboom & Cramer, 2013). The networks computed were therefore fully connected. Networks were drawn using the default Fruchterman-Reingold force-directed layout algorithm (Fruchterman & Reingold, 1991), in which symptoms with stronger and more numerous connections are positioned closer together and more centrally within the network. We analysed the depression-anxiety networks for the two age groups, as described below. All analyses were conducted using NetworkX package (Hagberg et al., 2008) in Python version 3.11.5.

### Network analyses

#### Community detection

First, we used the Louvain community detection algorithm to identify clusters or communities of highly interconnected symptoms within the network (Blondel et al., 2008). Communities represent groups of symptoms that are more strongly associated with each other than with symptoms in other communities. The Louvain method finds the “optimal” number of communities in a network, which maximises modularity (highest within-community connections and lowest between-community connections). The best fit community structure is fully data-driven, and does not require any a priori constraints. We next conducted several analyses on these communities, as described below. However, to confirm that the results of these analyses were not driven by age-related differences in community clustering, we conducted similar analyses on the a priori symptom assignment of the HADS, with one community for anxiety symptoms (HA1-HA7) and another community for depressive symptoms (HD1-HD7).

#### Within- and between-community connections

Once communities were identified, we examined the strength of connections within- and between-community. We calculated the mean within-community connection strength as the average strength of edge weights for within-community edges, and mean between-community connection strength as the average strength of edge weights for edges connecting symptoms between communities. These metrics were calculated for each community and for each age group network, and we compared them across communities and age using permutation tests, as done in previous research (Bekhuis et al., 2016). Specifically, in each permutation test, we randomly shuffled the age group labels (or community assignments in the case of community comparison) and recalculated the difference in mean edge weight between the two groups being compared. This process was repeated 5,000 times to generate a null distribution of differences under the assumption of no true difference between the groups. The p-value was then calculated as the proportion of permutations where the absolute difference in mean edge weight was equal to, or greater than, the observed absolute difference in the original data.

#### Strength centrality and bridge centrality

We calculated metrics of strength centrality and bridge centrality for each symptom across the network. First, centrality measures reflect how strongly connected a particular symptom is to all other symptoms across the whole network. We focused on node strength centrality, a widely-used and reliable metric defined as the sum of all connections a given symptom has with other nodes (Fried et al., 2016). Strength centrality is known to align well with other centrality metrics and is often considered the most suitable indicator of symptom centrality in psychopathology networks, due to its ability to capture the overall influence of a symptom within the network (Park & Kim, 2020). Other centrality measures, such as closeness and betweenness were not computed, as they have no clear meaning for full graphs, i.e., fully connected graphs, such as those used in our study (Bringmann et al., 2019; Jones et al., 2021). Second, bridge strength was calculated, which assesses the importance of a symptom in connecting different communities within the network (Jones et al., 2021). Specifically, bridge strength for a node was calculated as the sum of the weights of its edges that connect to nodes in other communities. This differs from strength centrality, which considers connections to all nodes regardless of community membership. We used Spearman correlation analysis to examine the consistency of strength centrality and bridge strength patterns between young and older adult groups (De Winter et al., 2016).

## RESULTS

Our cohort comprised 786 participants. Key summary statistics, including age, sex, mean total HADS-D, and mean total HADS-A scores across age groups, are summarized in Table S1. While no significant differences were found for sex or overall depressive symptom severity (*t*(408.20) = 0.71, *p* = 0.475), overall anxiety symptom severity was significantly higher in younger adults (*M* = 8.25, *SD* = 3.56) compared to older adults (*M* = 5.70, *SD* = 3.55), *t*(433.07) = 9.16, *p* < 0.001.

Before turning to the network analyses, we first examined the severity of each depressive and anxiety symptom across age groups (Table 1), by conducting pairwise comparisons for each item and correcting for multiple comparisons using the false discovery rate (FDR). Younger adults generally reported significantly higher severity in most anxiety symptoms, while differences in depressive symptoms were more mixed. Older adults reported higher severity in specific depressive symptoms related to fatigue (HD4), decreased motivation (HD6) and sustained enjoyment (HD1), while younger adults reported higher severity in symptoms related to depressed mood (HD3) and anhedonia (HD7). However, our focus was on the structure of the relationship between these symptoms, for which we employed a symptom network approach.

**Table 1.** Depressive and anxiety item comparison between younger and older adults. The table presents the mean (and SD) for each HADS item in the younger and older adult groups. *p*-values were obtained from independent t-tests comparing the means between the two groups and were corrected for multiple comparisons using the false discovery rate (FDR).

| Item | Young Mean (SD) | Old Mean (SD) | U-Statistics | p-value (FDR Corrected) |
| --- | --- | --- | --- | --- |
| Enjoyment (HD1) | 0.46 (0.67) | 0.61 (0.78) | 415579.0 | <0.001 |
| Laughter (HD2) | 0.26 (0.50) | 0.27 (0.54) | 466113.0 | 0.89 |
| Cheerful Feeling (HD3) | 0.35 (0.54) | 0.27 (0.52) | 500249.0 | <0.001 |
| Slowed Down (HD4) | 0.70 (0.72) | 1.36 (0.93) | 282006.0 | <0.001 |
| Loss of interest in Appearance (HD5) | 0.50 (0.74) | 0.47 (0.71) | 471670.0 | 0.52 |
| Look Forward to Things (HD6) | 0.37 (0.65) | 0.53 (0.72) | 408737.5 | <0.001 |
| Enjoyment of Book/Media (HD7) | 0.28 (0.63) | 0.19 (0.52) | 492970.0 | <0.001 |
| Feeling of Tension (HA1) | 1.06 (0.65) | 0.78 (0.62) | 566009.5 | <0.001 |
| Frightened Feeling (HA2) | 0.69 (0.81) | 0.53 (0.74) | 513962.0 | <0.001 |
| Worrying Thoughts (HA3) | 0.98 (0.87) | 0.76 (0.80) | 531450.0 | <0.001 |
| Relaxed Feeling (HA4) | 0.84 (0.68) | 0.60 (0.63) | 551580.0 | <0.001 |
| Butterflies in Stomach (HA5) | 0.61 (0.64) | 0.43 (0.59) | 538413.0 | <0.001 |
| Restless Feeling (HA6) | 1.21 (0.90) | 0.92 (0.82) | 547494.5 | <0.001 |
| Feeling of Panic (HA7) | 0.54 (0.67) | 0.49 (0.65) | 483132.0 | 0.09 |

### Depression-anxiety symptom networks and communities

After computing depression-anxiety symptom network for each age group, we conducted a community detection analysis to identify distinct clusters or communities of symptoms within the networks (Fig. 1). In the network of younger adults, two communities were detected. One community predominantly included anxiety-related items (HA1-HA3, HA5, HA7), while the other community included depressive items (HD1-HD7) along with the anxiety items “relaxed feeling” (HA4) and “restless feeling” (HA6). In older adults, the network showed a similar community structure. There were again two communities, with a small overlap between depressive and anxiety items. One community predominantly included anxiety-related items (HA1-HA3 and HA5-HA7), while the other community included depression-related items (HD1-HD7) and the anxiety item “relaxed feeling” (HA4). This suggests that symptom communities remained largely similar across age groups, with two predominant depression- and anxiety-related symptom communities.

**Figure 1.**
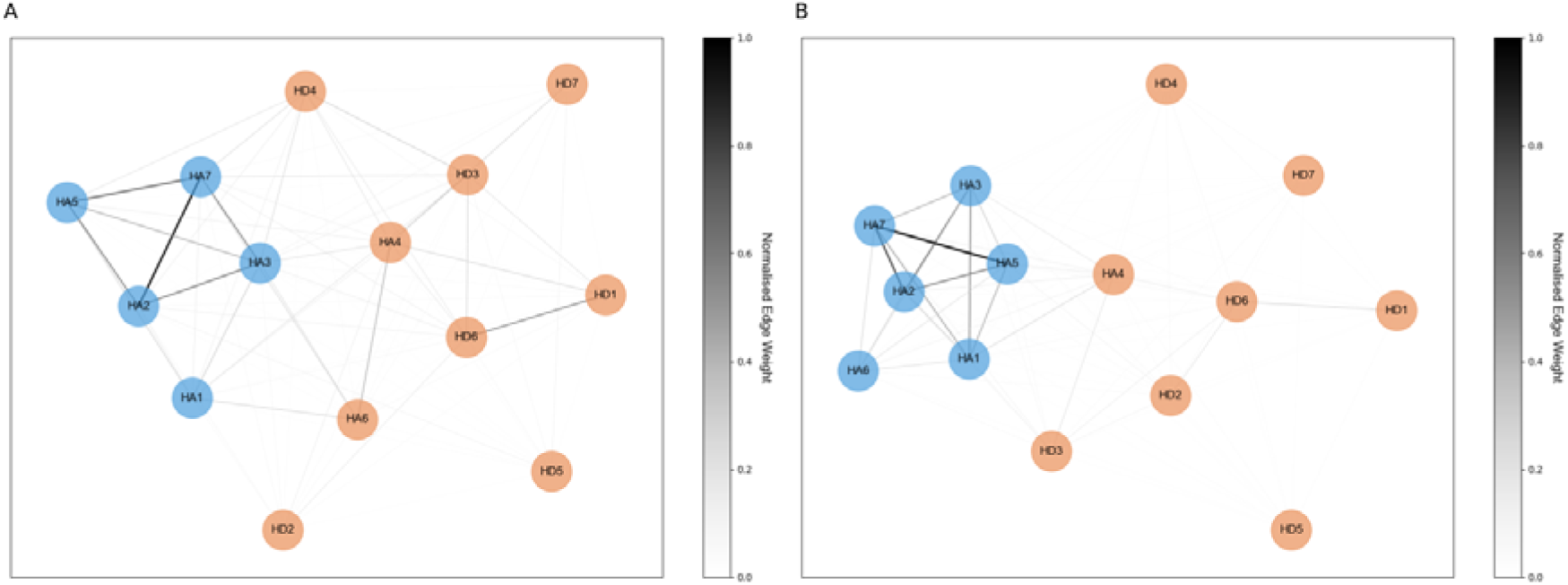
Depression-anxiety symptom networks in younger and older adults. (A) Network constructed using mutual information (MI) between HADS questionnaire items in younger adults, with edges representing the magnitude of MI associations between items. Edge thickness and colour (as indicated in the colour bar) indicate normalized MI. Node colours represent communities detected by the Louvain algorithm. (Blue shade: anxiety community, orange shade: depression community). (B) Same as (A) but for older adults.

We analysed the network based on these communities which were detected in a data-driven manner. However, to confirm that any observed group differences in these analyses are not due to the small differences in symptom clustering into communities between age groups, we also performed these analyses on the symptom clusters according to their assignment in the HADS, i.e., one community for anxiety items (HA) and another community for depression items (HD). The results were similar to those described below (see Supplementary Results).

### Within- and Between-Community Connectivity

We examined both within-community connectivity (strength of connections within each community) and between-community connectivity (strength of connections between communities) for both age groups (Fig. 2). Overall, the anxiety-related symptom community exhibited higher within-community connectivity compared to the depression-related symptom community across both age groups. Importantly, there was an age-related increase in within-community connectivity specifically for the anxiety-related symptom community, with significantly higher connectivity in older adults (*M* = 0.519) compared to young adults (*M* = 0.423; *p* < 0.001, permutation test). In contrast, within-community connectivity in the depression-related symptom community significantly decreased with age, showing lower connectivity in older adults (*M* = 0.160) compared to young adults (*M* = 0.298; *p* < 0.001, permutation test).

**Figure 2.**
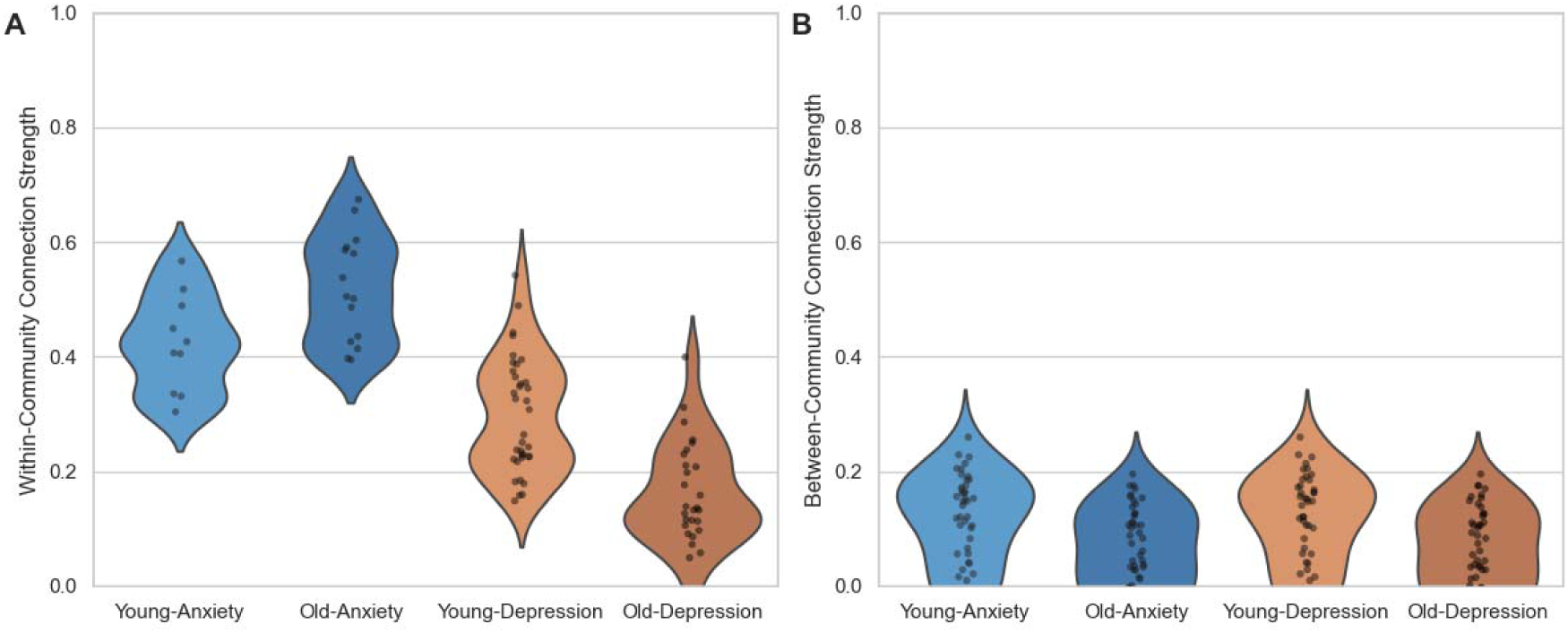
Age-related differences in within- and between-community connectivity. (A) Violin plots displaying the distribution of within-community connection strength for each symptom within the depression (orange shade) and anxiety (blue shade) symptom communities, for both young (lighter shades) and older adults (darker shade). (B) Same as (A) but for between-community connections.

For between-community connections, across both communities, we found a significant age-related decline in between-community connectivity (*p* < 0.001, permutation test) in older adults (*M* = 0.0625) compared to young adults (*M* = 0.1112). This pattern was consistent for both the depression and anxiety communities.

### Strength centrality and bridge strength

We examined both strength centrality and bridge strength measures across age groups. In terms of strength centrality (Fig. 3A), item HA7 (“panic”) exhibited the highest strength centrality in both young and older adult groups. Moreover, across items, strength centrality pattern appeared similar between age groups. A (ranked) correlation analysis showed a near-perfect alignment in symptom centrality ranking between age groups (ρ = 0.999, *p* < 0.001), suggesting that the influence of individual symptoms within the whole network was largely stable with age.

**Figure 3.**
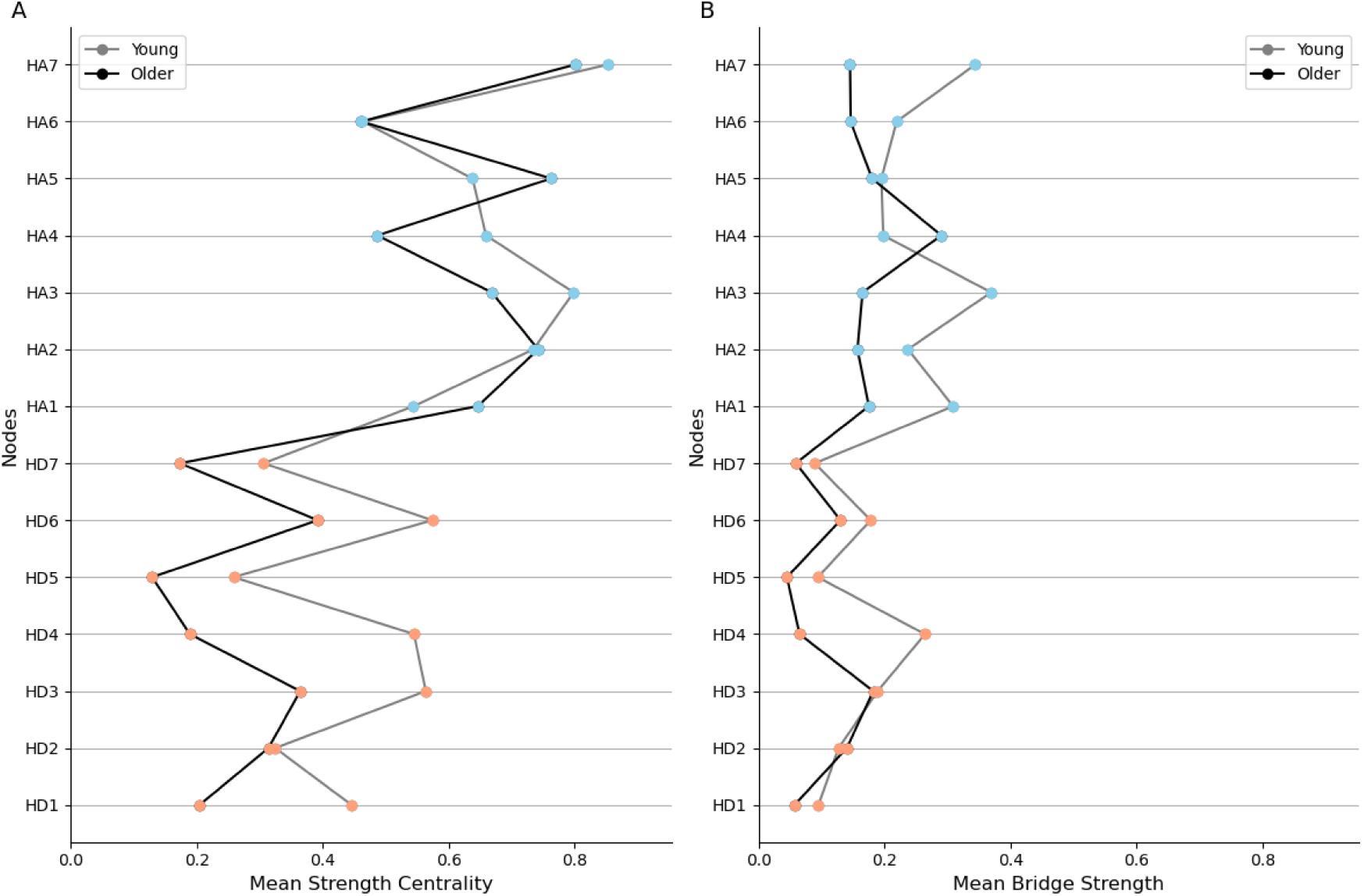
Comparison of centrality measures between age groups. (A) Strength centrality, representing the sum of connection weights for each symptom, is displayed for both young (gray line) and older adults (black line). Nodes are colored according to their classification in the Hospital Depression and anxiety Scale (HADS), with anxiety symptoms (HA) in blue and depressive symptoms (HD) in orange. (B) Bridge strength centrality, which quantifies the importance of a symptom in connecting the depression and anxiety communities, is shown for young and older adults. Each node is represented by a gray line connecting its bridge strength values in the young (gray line) and older adults (black line) groups. Node colors correspond to their classification in the HADS, as in (A).

In contrast to the consistent yet reduced centrality measures for the depressive symptom community, bridge strength pattern differed with age (Fig. 3B). Specifically, HA3 (“rumination”) exhibited the highest bridge strength in young adults, whereas HA4 (“restlessness”) exhibited the highest bridge strength in older adults. Spearman correlation analysis showed a moderate positive correlation between the rank of bridge strength scores in young and older adults (ρ = 0.51, *p* = 0.064), indicating a weak consistency in the bridging role of nodes across age groups.

## DISCUSSION

Our study employed network analyses to examine the complex structure of depressive and anxiety symptoms in the general population across the lifespan. Our principal findings include an overall stability with age for network organization and centrality; an age-related increase in within-community connection strength for anxiety but decrease in within-community connection for depression; age-specific bridging symptoms, with “rumination” and “restlessness” as the most prominent bridging symptoms in young and older adults, respectively. Together, the results suggest that anxiety and depression symptom networks evolve with age, which has important implications for age-specific tailoring of treatment.

Direct comparisons of symptom severity showed age-related differences. Younger individuals with subclinical depressive symptoms tended to report more severe anxiety symptoms compared to older adults. This observation aligns with previous research highlighting a stronger association between depression and anxiety in younger populations (Beekman et al., 2000, 2000). However, some studies have reported contrasting findings, potentially due to age-related biases in assessment tools or the influence of other age-dependent factors, such as physical health comorbidities, cognitive decline, social isolation, and bereavement (Jorm, 2000; Wuthrich et al., 2015).

In terms of symptom networks, symptom communities remained consistent across age groups, and largely mapped onto depressive and anxiety symptoms of the HADS. This suggests that the core organization of psychopathological networks might be preserved across different populations and contexts (Boschloo et al., 2015).

Interestingly, some anxiety symptoms were clustered into the depressive symptom community and vice versa for some depressive symptoms. Specifically, “difficulties in relaxation” and “restless feelings” were clustered into the depression-related community. This suggests a potential blurring of the boundaries between depression and anxiety, particularly in relation to somatic symptoms. Moreover, previous studies using the HADS have found a three-dimensional model, which includes a “psychomotor agitation” dimension that is strongly correlated with depression (Caci et al., 2003; Friedman et al., 2001). While our network analysis did not explicitly identify three distinct communities, the closer association of certain anxiety symptoms with the depressive community suggests a potential overlap between these dimensions.

Examining the connections within and between communities, a notable result was the age-related decline in the connectivity of the depressive symptom community, while connectivity in the anxiety community significantly increased with age. This divergence suggests age-specific changes in how these symptom domains interact and manifest. The decline in depressive symptom connectivity may reflect greater heterogeneity in reported depressive symptoms among older adults, potentially driven by comorbid medical conditions and social isolation (Jellinger, 2023; Korten et al., 2012). From a clinical perspective, this age-related change might imply not only that depressive symptoms are less likely to cluster together in older adults, but also that targeting a single or core symptom might be less effective in alleviating the broader depressive experience in this age group. Evidence for smaller clinical efficacy of depression treatment in older adults is widely reported (Alexopoulos, 2019; Schaakxs et al., 2018), calling for more individualized treatment approaches. In contrast, the increased within-community connectivity of anxiety symptoms suggests a more consolidated anxiety presentation in older adults, which could reflect heightened interrelatedness or a tendency for these symptoms to co-occur more strongly with age (Ehring et al., 2011).

Between-community connectivity declined significantly with age, suggesting a diminishing interplay between depression and anxiety symptoms. This reduction was particularly pronounced for connections involving anxiety symptoms. This finding might reflect age-related changes in cognitive-emotional processing, including shifts in attention, memory, and emotional regulation, which can alter the interplay between affective symptoms (Charles & Carstensen, 2010; Mather & Knight, 2005). Knight and Durbin (Knight & Durbin, 2015) further propose that older adults may develop more adaptive coping mechanisms, such as prioritizing emotionally meaningful goals and reducing engagement in ruminative thinking, which may contribute to a reduced coupling of depression and anxiety symptoms. As cognitive control processes change with age, particularly in the context of emotion regulation and thought patterns, the distinction between depression and anxiety may become more pronounced. Future studies should investigate how cognitive changes, particularly in executive functions and emotional regulation, contribute to these age-related alterations in symptom connectivity, and how these insights could guide age-specific therapeutic strategies.

The analysis of symptom centrality again revealed some age-related consistencies but also discrepancies in the structure of depression-anxiety symptom network. In both age groups, anxiety symptoms, and particularly “panic”, were more central within the symptom network compared to depressive symptoms (Park & Kim, 2020), echoing some who suggest anxiety symptoms play a prominent role in the overall experience of distress relative to depressive symptoms (Kessler et al., 2006). Importantly, we observed an age-related difference in bridging symptoms, with rumination emerging as the strongest bridge symptom in younger adult, but restlessness emerging as the strongest bridge symptom in older adults. This suggests a potential transition in the primary mechanisms linking depression and anxiety across the lifespan, with a possible shift from cognitive (rumination) to somatic (restlessness) manifestations of depression-anxiety in older age. Supporting this interpretation, evidence suggests that depression and anxiety can manifest differently in older adults, with a greater emphasis on somatic complaints and physical symptoms (Balsis & Cully, 2008; Wolitzky-Taylor et al., 2010).

### Strengths and limitations

Our study has several significant strengths. We used validated scales (HADS) within a large, population-derived cohort, encompassing a wide range of ages. Participants were diverse and more representative of the general population (Shafto et al., 2014), with balanced sociodemographic characteristics and a similar number of individuals in each age decile. Networks were computed using mutual information, which is a robust statistical method for detecting non-linear relationships between variables. However, our study also has several limitations. First, the cross-sectional design precludes the establishment of causal relationships or the assessment of changes in depressive and anxiety symptoms over time. Second, the reliance on self-report questionnaires for symptom assessment, rather than clinician assessment, introduces the potential for biases and reporting inaccuracies. Third, the examination of non-clinical general population, while offering valuable insights into ‘real-world’ symptom patterns in the general community, limits the generalizability of our findings to clinically diagnosed populations with depression and anxiety. Furthermore, the HADS, although a widely used screening tool, does not encompass the full spectrum of depressive and anxiety manifestations as outlined in diagnostic manuals.

### Conclusion

We found evidence for both stability and age-related differences in anxiety-depression symptom network. The core network structure and the centrality of certain symptoms remained consistent with group. However, in older compared to young adults, differences emerged specifically for depressive symptoms, where there was a transition from cognitive to somatic bridging symptoms, and a decline in within-community connectivity. These results suggest that while anxiety symptoms maintain their relationship with age, depressive symptoms are more heterogeneous, which may suggest different underlying mechanisms. These findings underscore the need for tailored clinical approaches that recognize both the consistent and evolving nature of depression and anxiety symptoms across the lifespan. Future research would further investigate the mechanisms underlying these age-related differences and their implications for treatment.

## Data Availability

All data produced in the present study are available upon reasonable request to the authors

## Supplementary Results

This section presents the results of analyses conducted using a priori-defined communities based on HADS symptom assignment, confirming that the main findings were not solely driven by age-related differences in community clustering identified using the Louvain method.

**Table S1:** This table provides demographic and clinical characteristics of the study population, including sex, age, and mean Hospital Depression and anxiety Scale-Anxiety (HADS-A) and Hospital Depression and anxiety Scale-Depression (HADS-D) scores for young and older adults.

**Figure S1:** This figure displays violin plots comparing age-related differences in within-and between-community connectivity for depression and anxiety symptom communities, based on a priori-defined communities from the HADS symptom assignment.

