## Supplementary Material for "Depression and Anxiety Symptom Networks Across the Lifespan"

### SUPPLEMENTARY INFORMATION

**Supplementary Results**

To address potential age group differences in symptom clustering, we re-ran all analyses reported in the main text using *a priori*-defined communities based on HADS symptom assignment—one community for all anxiety symptoms (HA1-HA7) and another community for all depression symptoms (HD1-HD7). This analysis confirms that the main results were not solely driven by the small age-related differences in community clustering identified using the Louvain method.

*Within-Community Connectivity:* We first compared the average within-community connection strength for the depression and anxiety communities across both age groups. Consistent with the results from communities detected using the Louvain method, the anxiety community exhibited higher average within-community connection strength compared to the depression community across both age groups (Young-Anxiety: $M = 0.447$, Older-Anxiety: $M = 0.527$; Young-Depression: $M = 0.209$, Older-Depression: $M = 0.127$). Significant age-related differences were observed; specifically, within-community connection strength for the depression community declined significantly with age ($p < 0.001$, permutation test), while the within-community connection strength in the anxiety community was also significantly higher in older adults compared to younger adults ($p = 0.0018$, permutation test).

*Between-Community Connectivity:* We examined between-community connectivity. As expected in the *a priori* analysis, between-community connectivity was equal for the depression and anxiety communities within each age group; however, we observed a significant age-related decline, with lower connectivity in older adults (Anxiety/Depression: $M = 0.0625$,

*SD* = 0.0451) compared to young adults (Anxiety/Depression: *M* = 0.1112, *SD* = 0.0776; $p < 0.001$, permutation test).

*Consistency with Main Results:* Overall, these results align with the main findings using communities detected with the Louvain algorithm, suggesting that observed differences in connectivity patterns are not solely driven by age-related changes in community clustering. This analysis further supports that age-related differences in connectivity patterns are robust and not dependent on the method of community detection.

**Table S1. Demographic and clinical characteristics of the study population.** The table provides the distribution of sex (number of male/female participants), mean age (and standard deviation), mean Hospital Depression and anxiety Scale-Anxiety (HADS-A) score (and standard deviation), and mean Hospital Depression and anxiety Scale-Depression (HADS-D) score (and standard deviation) for the study population. t-values and p-values represent the results of independent t-tests comparing these measures between young and older adults.

| Variable | Young adults | Older adults | t-value | p-value |
|---|---|---|---|---|
| **Sex M/F** | 107/125 | 223 /231 | - | 0.13 |
| **Age** | 33.42 (7.05) | 81.29 (7.10) | - | - |
| **HADS-D** | 6.32 (2.47) | 6.19 (2.31) | 0.62 | 0.46 |
| **HADS-A** | 8.25 (3.56) | 5.70 (3.55) | -7.31 | <0.001 |

**Figure S1. A priori comparison of age-related differences in within- and between-community connectivity.** (A) Violin plots displaying the distribution of within-community connection strength for each symptom within the depression (orange shade) and anxiety (blue shade) symptom communities, for both young (lighter shades) and older adults (darker shade). (B) Same as (A) but for between-community connections.

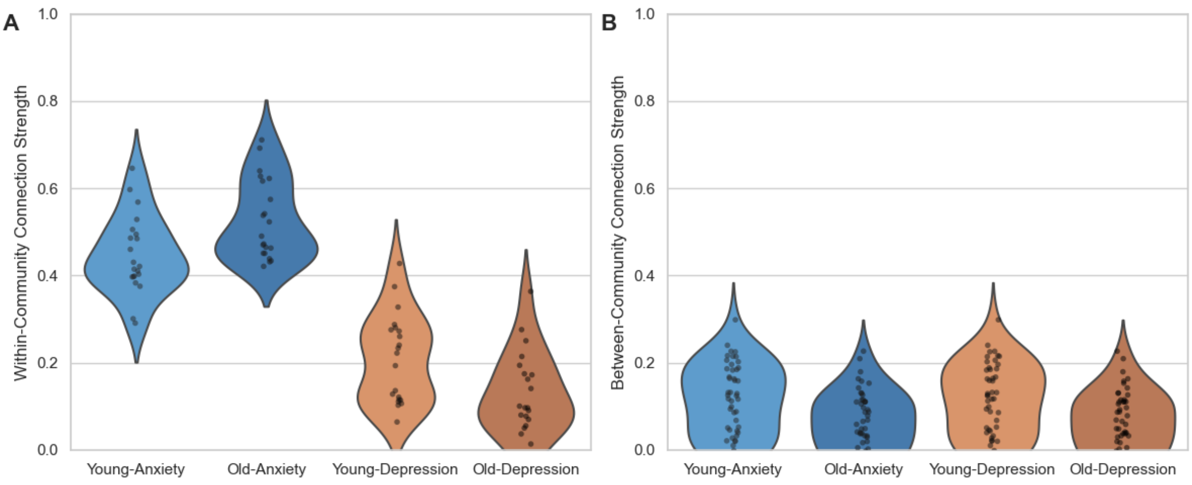
